# Progression, recovery and fatality in patients with SARS-CoV-2 related pneumonia in Wuhan, China: a single-centered, retrospective, observational study

**DOI:** 10.1101/2020.05.12.20099739

**Authors:** Yirong Lu, Qingquan Lv, Xiping Wu, Tian Hu, Kai Wang, Yumei Liu, Yuhai Hu, Lan Yu, Hexuan Fei, Zheng Ba, Xiaohua Lin, Hua Wang

## Abstract

**Objectives:** To determine the case fatality rates and death risk factors.

**Design:** Retrospective case series.

**Setting:** A COVID-19 ward of a secondary Hospital in Wuhan, China.

**Participants:** Consecutively hospitalized COVID-19 patients between Jan 3, 2020 and Feb 27, 2020. Outcomes were followed up to discharge or death.

**Results:** Of 121 patients included, 66 (54.6%) were males. The median age was 59 (IQR: 46 to 67) years, and hypertension (33 patients; 27.3%) the leading comorbidity. Lymphopenia (83 of 115 patients; 72.2%) frequently occurred and then normalized on day 4 (IQR: 3 to 6) after admission in the survivors, with lung lesion absorbed gradually on day 8 (IQR: 6 to10) after onset (33 of 57 patients; 57.9%). The real-time polymerase chain reaction (RT-PCR) assays for SARS-CoV-2 were positive in 78 (78/108; 72.2%) patients, and a false-negative RT-PCR occurred in 15 (13.9%) patients. Hypoxemia occurred in 94 (94/117; 80.3%) patients, and supplemental oxygen was given in 88 (72.7%) patients, and mon-invasive or invasive ventilation in 20 (16.5%) cases. Corticosteroid use might link to death. The case fatality rates were 4.4% (one of 23 patients), 29.3% (12/41), 22.8% (13/57) or 45% (9/20) for patients with moderate, severe, critical illness or on ventilator. The length of hospital stay was 14 (IQR: 10 to 20) days, and selfcare ability worsened in 21 patients (21/66; 31.8%) cases. Patients over 60 years were most likely to have poorer outcomes, and increasing in age by one-year increased risk for death by 18% (CI: 1.04-1.32).

**Conclusions:** In management of patients with SARS-CoV-2 pneumonia, especially the elderly with hypertension, close monitoring and appropriate supportive treatment should be taken earlier and aggressively to prevent from developing severe or critical illness. Corticosteroid use might link to death. Repeated RT-PCR tests or novel detection methods for SARS-CoV-2 should be adopted to improve diagnostic efficiency.

**Strengths and limitations of this study:**

➢ Eight case series reported mortality of 6.2% to 61.5% in COVID-19 patients in Wuhan, China. However, outcomes were inadequately followed and the risk factors for death unrevealed.
➢ The case fatality rates were 4.4%, 29.3%, 22.8% or 45% for patients with moderate, severe, critical illness or on ventilator.
➢ Age was the independent factor for death, and an increase by one-year increased risk for death by 18% (odds ratio: 1.18; 95% CI: 1.04-1.32; *P* < .01).
➢ Case fatality rates calculated might be affected by small patient subset size and non-prospective data collection.

## INTRODUCTION

In late December 2019, a cluster of atypical pneumonia of unknown cause was initially noticed and reported in Wuhan, Hubei Province, China.^1^ On Jan 7, 2020, a bat SARS-like coronavirus strain was identified from bronchoalveolar lavage fluid samples in three local patients by real-time reverse transcription polymerase chain reaction (RT-PCR) assay, and the whole genome was sequenced and shared with the WHO.^2^ By far, the novel coronavirus had caused severe infections and global circulation in humans,^3^ and known as a member of beta-coronavirus of the subgenus Sarbecovirus, with at least 79.5% of genetic sequence with SARS-CoV and 96.2% of homology to the coronavirus strain BatCov RaTG13.^4^ Then, this SARS-like coronavirus was named as SARS-CoV-2 by the International Committee for Taxonomy of Virus (ICTV), and the related infection as COVID-19 by the WHO on Feb 11, 2020.^5 6^

The COVID-19 outbreak in Wuhan could be traced back to Dec 1, 2019,^7^ while a local resident was documented with respiratory symptoms and then confirmed as SARS-CoV-2 pneumonia, though any epidemical link was not found between the patient and other cases. From then on, the epidemic had rapidly grown, with a reported number of 41 suspected cases in Wuhan on Jan 3, 2020,^8^ and then 1284 confirmed cases on Jan 24, 2020, the Eve of Chinese Lunar New Year. Thereafter, the curve of reported case was peaked and flattened on Feb 18, 2020 or so, with a total of 50, 633 confirmed cases in Wuhan and Hubei Province, 7, 253 from the other 30 provinces and regions, China. Meanwhile, 871 exported cases were reported from 25 countries across the world, a precursor of global pandemic.^8 9^

The figure reported was just the tip of an iceberg, victims might be far beyond the known.^9^ On Jan 23, 2020, traffic suspension was announced by Wuhan authorities, with community containment, to reduce movement of people and block virus transmission. As of Feb 28, 2020, forty-five designated and 12 temporary hospitals with more than 50,000 beds in Wuhan were urgently reconstructed, and more than 42,000 healthcare workers deployed across other areas of China, in response to the SARS-CoV-2 outbreak. Since Jan 26, 2020, as one of the firstly arrived medical assistance teams in Wuhan, we had been working at a COVID-19 ward in Hankou Hospital, one of the three firstly designated COVID-19 hospitals on Jan 3, 2020. Herein, given that the case fatality rates and death risk factors were not fully known for patients with SARS-CoV-2 pneumonia in the prodromal stage of the epidemic, and the knowledge pertaining to the progression of the disease was limited in the published cohort studies,^7 10-14^ we reported a cases series with SARS-CoV-2 associated pneumonia, most of whom were severely and critically ill and treated in our COVID-19 ward, to describe the natural process of the disease and risk factors for critical illness or death.

### Methods

#### Study design and participants

This project was a single-centered, retrospective, observational cohort study, conducted at a COVID-19 ward in a secondary hospital, Wuhan, China. Patients admitted between Jan 3, 2020 and Feb 27, 2020 were screened for eligibility and those with a diagnosis of laboratory-confirmed or suspected SARS-CoV-2 pneumonia were included. Outcomes were followed up to hospital discharge or death.

Ethics approval was obtained from the Hankou Hospital Medical Research Ethics Committee (No. HKYY-2020-028), and written informed consent was waived for this retrospective study. In addition, the Strengthening the Reporting of Observational Studies in Epidemiology (STROBE) statement: guidelines for reporting observational studies was followed.^15^

#### Laboratory measurements

Serum biochemical tests were completed on admission and repeated according to the attending doctor’s decisions, including complete blood count, coagulation profile, bilirubin, creatinine, cardiac troponin, electrolytes and procalcitonin. Serum immunoglobulin M (IgM) and IgG were detected by immunogold labelling technique to exclude other pathogens, e.g., influenza A and B viruses, adenovirus, respiratory syncytial virus and parainfluenza virus. Sputum culture was also conducted on admission to identify possible causative bacteria or fungi.

RT-PCR assay for SARS-CoV-2 nucleic acid on nasopharyngeal swab was not available until late in the study, and was performed in the Wuhan Center for Disease Control and Prevention in accordance with the Recommendation by the National Institute for Viral Disease Control and Prevention (China).^16^ Meanwhile, we also detected quantitively serum IgM and IgG antibodies specific for SARS-CoV-2 in patients who were contemporarily hospitalized and remained negative RT-PCR testing, using microfluidic chemiluminescence immunoassay and an immune analyzer (MF07, Huamai, Co. Ltd. Shenzhen, CN). The antibody diagnostic test kit was provided by the same company, and detection was performed by a qualified technician (Y Hu), according to the product instruction, and a cutoff value of more than 1.0 absorbance unit per milliliter was considered positive.

Two experienced radiologists (LY and HF) who were blinded to the study independently reviewed the chest CT images through the Picture Archiving and Communication Systems (PACS, DJ HealthUnion Systems Corporation, Shanghai, CN), to delineate the radiological features and changes over time. Discrepancies were resolved by consensus.

#### Management of patients

Oseltamivir was given empirically for antiviral therapy, and antibiotics prescribed alone or in combination to prevent secondary bacterial infection. Methylprednisolone and immunoglobulin were also administered intravenously for 7-10 days, based on the degree of severity of the disease. Moreover, thymopentin and cordyceps sinensis (herb medicine) were given for repletion of lymphocytes. Due to the inadequate supply of central oxygen, supplemental oxygen was given largely by nasal cannula with an average flow rate of 3 L/min, and oxygen tanks, oxygenators plus mask with or without a reservoir were adopted to increase the fraction of inspired oxygen concentration (FiO_2_). High flow nasal cannula and noninvasive ventilation were instituted in critically ill patients, with a FiO_2_ of 50%-60%, while mechanical ventilation and extracorporeal membrane oxygenation (ECMO) could hardly be put into practice.

#### Definition

SARS-CoV-2 related pneumonia referred to lung infection of suspected or confirmed SARS-CoV-2 origin. The diagnostic criteria for suspected SARS-CoV-2 related pneumonia were consistent with the Draft Protocol for Diagnosis and Treatment of SARS-CoV-2 Pneumonia (in Chinese, 6th edition).^17^ (1) within 14 days before symptom onset, the patient had any of the followings: residence history in Wuhan or the vicinity, or travel history to an area with documented SARS-CoV-2 infection; close contact with a COVID-19 patient; close contact with a person from a community where SARS-CoV-2 infections have been reported; clustered onset. (2) Present at least two of the followings: fever or respiratory symptoms; normal or reduced white blood cell counts, or lymphopenia in early onset; chest CT scan showed multifocal mottling and interstitial changes in lung periphery at the early-stage of the disease, then progressing to bilateral multifocal ground-glass opacities or infiltrations, and to massive consolidations in the later stage; (4) exclusion of influenza type A and B, adenovirus, respiratory syncytial virus, bacterial and other infectious respiratory diseases. The definite diagnosis for SARS-CoV-2 pneumonia was confirmed if the followings were met: (1) presence of epidemiological and clinical features as described above; (2) positive RT-PCR for SARS-CoV-2 nucleic acid on nasopharyngeal swab, or viral genome sequence obtained was highly homologous with those of SARS-CoV-2.

Severe pneumonia was determined if any of the followings were met:^17^ (1) shortness of breath, with respiratory rate ≥ 30 breaths per minute; (2) a pulse oxygen saturation ≤ 93% at rest, or a ratio of partial pressure of arterial oxygen to the fraction of inspired oxygen (PaO_2_/FiO_2_) ≤ 300 mm Hg (1 mm Hg = 0.133 kPa); (3) lung lesion extended ≥ 50% in radiographic images within 24-48 hours. Moreover, the critical illness was determined if any of the followings was documented: (1) respiratory failure requiring mechanical ventilation; (2) shock; (3) referral to the intensive care unit (ICU) with other organ dysfunction. The criteria for hospital discharge were (1) normal body temperature lasting for more than 3 days; (2) resolution of symptoms; (3) absorption of the lesions on chest CT image, and (4) consecutive RT-PCR testing negative for SARS-CoV-2 at least at a 1-day interval.

The date of illness onset was defined as the day when the symptom was noticed. Acute respiratory distress syndrome (ARDS) was defined in accordance with the Berlin definition,^18^ Acute kidney injury with the Kidney Disease: Improving Global Outcomes definition,^19^ Shock with the Surviving Sepsis Campaign definition,^20^ and acute liver injury with the Guidelines for the Management of Adult Acute and Acute-on-Chronic Liver Failure in the ICU.^21^ The cardiac injury was diagnosed if the serum level of cardiac troponin I was above the upper limit of the normal range or new abnormalities presented in electrocardiography and echocardiography.^10^

#### Data acquisition

Clinical and hospital records were reviewed, and data were collected and filled in a printed Case Report Form for SARS-CoV-2 outbreak (nCoV CRF), shared by the International Severe Acute Respiratory and Emerging Infection Consortium (ISARIC). The nCoV CRF consists of core and daily sections, and the core CRF section records epidemiological factors (e.g., a history of exposure, close contact with a COVID-19 patient, clustering), demographics (e.g., ethnics, gender, estimated age, professional, comorbidities and risk factors). The daily CRF documents the worst values per day during the patient’s hospitalization, including symptoms, vital signs and laboratory findings. Besides, medicine and respiratory support modalities, complications, outcomes and ability to selfcare at discharge were also recorded. Data unavailable from the medical records were obtained by direct communication with the patient or a proxy.

#### Statistical analysis

Discrete variables were presented by frequencies (n) and percentages (%), and continuous variables by mean (SD), or median (IQR). The analyses regarding different factors were based on non-missing data, and missing data were not imputed. Statistical analysis for continuous variables was performed by *t*-tests or one-way analysis of variance for normally distributed variables, or by Wilcoxon rank-sum tests for non-normally distributed ones. The Pearson chi-square test, Fisher exact test or Mann-Whitney U test was used, as appropriate, for categorical data. To determine variables associated with in-hospital mortality, covariates assumed to be associated with death at *P* ≤ 0.05 in univariate analyses were entered in a binary stepwise backward logistic regression model. Kaplan-Meier plot with log-rank test was used for survival data. Bivariate Cox proportional hazard ratio (HR) model was used to determine HRs and 95% CIs between individual factors for death. All statistical tests were two-sided, *P* ≤ 0.05 was considered statistically significant. Statistical analyses were performed with SPSS 13.0 software packages or R software (version 3.6.0, R Foundation for Statistical Computing).

#### Patient and public involvement

Patients or the public were not involved in the design, or conduct, or reporting, or dissemination plans of this research.

### Results

#### Enrollment and baseline characteristics

A total of 169 patients were screened and 121 were eligible for enrollment in the study (**Figure 1**). All patients were adults and males accounted for 66 (54.6%), and the median age was 59 years old (IQR: 46-67; range, 21-85 years). The outcomes were followed up from the illness onset to discharge or death, and the final date of follow-up was Mar 2, 2020.

**Figure 1.**
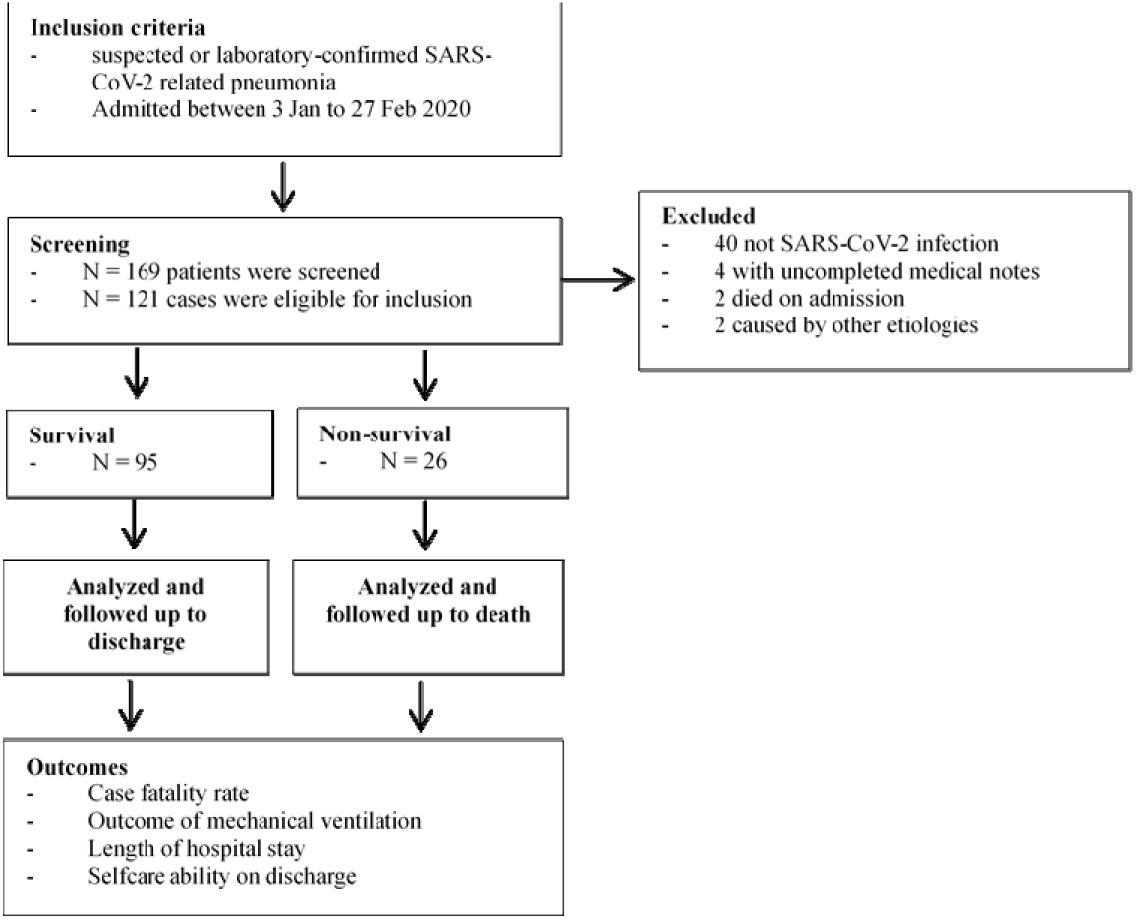
Study design and enrollment diagram.

Of 121 patients, 61 (50.4%) cases were retired, 20 (16.5%) workers, 13 (10.7%) company staff, four (3.3%) service personal including a bus driver and a cleaning worker, three (2.5%) teachers and student, and two (1.7%) doctors. Most patients (119; 98.4%) lived in Wuhan or the vicinity in 14 days before symptom onset, 12 (9.9%) had a history of exposure to the healthcare facilities, two (1.7%) to the Huanan Seafood Wholesale Market, nine (7.4%) had close contact to a COVID-19 persons, and six (5%) were familial clusters. Seventy (57.9%) cases had at least one or more underlying diseases, and hypertension (33 patients; 27.3%) was the leading comorbidity, followed by diabetes (25; 20.7%), liver disease (ten patients; 8.3%) including fatty liver and chronic hepatitis B, chronic cardiovascular disease (ten patients; 8.3%), chronic lung and kidney disease (five patients; 4.1%, respectively), and malignancy (four patients; 3.3%), (**Table 1**).

**Table 1.** Demographics, epidemiological, clinical and radiographic findings in survivors and non-survivors with SARS-CoV-2 related pneumonia.

|  | Total (n = 121) | Survivors (n = 95) | Non-Survivors (n = 26) | P Value<sup>a</sup> |
| --- | --- | --- | --- | --- |
| **Characteristics** |  |  |  |  |
| Median (IQR) age, years | 59 (46–67) | 58 (43–65) | 65.5 (56.8–73) | .002 |
| < 45 years | 28 (23.1) | 27 (28.4) | 1 (3.8) | .01 |
| 45–59 years | 35 (28.9) | 27 (28.4) | 8 (30.8) | .50 |
| ≥ 60 years | 58 (47.9) | 41 (43.2) | 17 (65.4) | .04 |
| Sex at birth |  |  |  |  |
| Male | 66 (54.6) | 51 (53.7) | 15 (57.7) | .45 |
| Female | 55 (45.5) | 44 (46.3) | 11 (42.3) | .45 |
| Body weight, median (IQR), kilogram | 65 (56–69) | 65 (58–70) | 64.5 (55–68.5) | .88 |
| Current smoker | 6 (5.0) | 5 (5.3) | 1 (3.8) | .62 |
| Professions |  |  |  |  |
| Retired | 61 (50.4) | 43 (45.3) | 18 (69.2) | .03 |
| Worker | 20 (16.5) | 17 (17.9) | 3 (11.5) | .44 |
| Company staff | 13 (10.7) | 13 (13.7) | 0 (0) | .07 |
| Service personal | 4 (3.3) | 4 (4.2) | 0 (0) | .58 |
| Freelancer | 4 (3.3) | 3 (3.2) | 1 (3.8) | > .99 |
| Teacher/student | 3 (2.5) | 3 (3.2) | 0 (0) | .58 |
| Healthcare staff | 2 (1.7) | 2 (2.1) | 0 (0) | > .99 |
| Others | 11 (9.1) | 10 (10.5) | 1 (3.8) | .45 |
| Epidemiology |  |  |  |  |
| Local resident | 118 (97.5) | 92 (96.8) | 26 (100) | 0.60 |
| Exposure to hospital | 12 (9.9) | 11 (11.6) | 1 (3.8) | 0.30 |
| Exposure to the seafood market | 2 (1.7) | 2 (2.1) | 0 (0) | > .99 |
| Close contact | 9 (7.4) | 9 (9.5) | 0 (0) | .20 |
| Clustering | 6 (5.0) | 6 (6.3) | 0 (0) | .34 |
| Others | 1 (0.8) | 1 (1.1) | 0 (0) | > .99 |
| Comorbidities |  |  |  |  |
| Hypertension | 33 (27.3) | 21 (22.1) | 12 (41.2) | .02 |
| Diabetes | 25 (20.7) | 18 (18.9) | 7 (26.9) | .37 |
| Liver disease | 10 (8.3) | 9 (9.5) | 1 (3.8) | .46 |
| Chronic heart disease | 10 (8.3) | 8 (8.4) | 3 (11.5) | .70 |
| Chronic lung disease | 5 (4.1) | 4 (4.2) | 1 (3.8) | .93 |
| Chronic kidney disease | 5 (4.1) | 4 (4.2) | 1 (3.8) | .93 |
| Malignancy | 4 (3.3) | 2 (2.1) | 2 (7.7) | .20 |
| Others | 15 (12.4) | 10 (10.5) | 5 (19.2) | .31 |
| **Symptom on admission** |  |  |  |  |
| Fever | 107 (88.4) | 84 (88.4) | 23 (88.5) | .65 |
| Cough | 69 (57) | 55 (57.9) | 14 (53.8) | .71 |
| Dry | 36 (52.2) | 30 (54.5) | 6 (42.9) | .40 |
| Haemoptysis | 4 (5.8) | 3 (5.5) | 1 (7.1) | > .99 |
| Dyspnea | 38 (31.4) | 26 (27.4) | 12 (46.2) | .07 |
| Median (IQR) time to dyspnea, day | 5 (3–9) | 5 (2–9) | 7 (3.3–9.8) | .52 |
| Fatigue | 33 (27.3) | 22 (23.2) | 11 (42.3) | .05 |
| Sore throat | 6 (5.0) | 3 (3.2) | 3 (11.5) | .11 |
| Diarrhoea/anorexia | 10 (8.3) | 8 (8.4) | 2 (7.7) | > .99 |
| Myalgia/arthralgia | 6 (5.0) | 6 (6.3) | 0 (0) | .34 |
| Headache/Confusion | 5 (4.1) | 5 (5.3) | 0 (0) | .36 |
| Others | 4 (5.8) | 2 (2.1) | 2 (7.7) | .20 |
| **Signs on admission** |  |  |  |  |
| Median (IQR) temperature at onset, °C, (36-37) | 38.5 (38~38.9) | 38.5 (38~39) | 38 (37.6~38.5) | .22 |
| 37-37.9 °C, n/N (%) | 22/93 (23.7) | 15/74 (20.3) | 7/19 (36.8) | .07 |
| 38-39 °C, n/N (%) | 49/93 (52.7) | 38/74 (51.4) | 11/19 (57.9) | .61 |
| > 39 °C, n/N (%) | 22/93 (23.7) | 21/74 (28.4) | 1/19 (5.3) | .07 |
| Median (IQR) heart rate, beat per minute, (60-100) | 86 (80-94) | 85 (80-98) | 88 (80-90) | .76 |
| 60-100, beats per minute | 101 (83.5) | 79 (83.2) | 22 (84.6) | .56 |
| > 100 beats per minute | 20 (16.5) | 16 (16.8) | 4 (15.4) | .56 |
| Median (IQR) respiratory rate, breath per minute, (16-20) | 20 (20-22) | 20 (19-22) | 20 (20-23) | .84 |
| ≥ 30 breaths per minute | 12 (9.9) | 10 (10.5) | 2 (7.7) | .36 |
| Median (IQR) MAP, mm Hg | 90 (85.7-95.2) | 90 (85.3-94.4) | 92.7 (86-96) | .18 |
| Median (IQR) PaO<sub>2</sub>/FiO<sub>2</sub> at admission, mm Hg, (400-500) | 227 (187.9-272.7) | 227 (193.9-272.7) | 203 (111.3-247.7) | .05 |
| ≤ 100 mm Hg, n/N (%) | 11/117 (9.4) | 5/91 (5.5) | 6/26 (23.1) | .03 |
| ≤ 200 mm Hg, n/N (%) | 32/117 (27.4) | 25/91 (27.5) | 7/26 (26.9) | .95 |
| ≤ 300 mm Hg, n/N (%) | 51/117 (43.6) | 39/91 (42.9) | 12/26 (75) | .77 |
| Chest CT findings, n/N (%) |  |  |  |  |
| Multifocal mottling | 80/116 (69.0) | 61/94 (64.9) | 19/22(86.4) | .07 |
| Ground-glass opacity | 73/116 (62.9) | 56/94 (59.6) | 17/22 (77.3) | .15 |
| Consolidation | 18/116 (15.5) | 17/94 (18.1) | 1/22 (4.5) | .19 |
| Parenchymal band | 8/116 (6.9) | 5/94 (5.3) | 3/22 (13.6) | .17 |
| Bilateral involvement | 105/116 (66.4) | 84/94 (89.4) | 21/22 (95.5) | .69 |
| Peripheral distribution | 87/116 (75.0) | 69/94 (73.4) | 18/22 (81.8) | .59 |
| Diffused infiltration | 22/116 (19.0) | 17/94 (18.1) | 5/22 (22.7) | .56 |
| Disease severity status |  |  |  |  |
| Moderate | 23 (19.0) | 22 (23.2) | 1 (3.9) | .04 |
| Severe | 41 (33.9) | 29 (30.5) | 12 (46.2) | .14 |
| Critical | 57 (47.1) | 44 (46.3) | 13 (5.0) | .74 |
| Median (IQR) time from onset to clinics, day | 3 (1~6) | 3 (1~6) | 4.5 (1.8~11.8) | .09 |
| Median (IQR) time from onset to admission, day | 7 (4~10) | 7 (4~9) | 7 (5~12) | .13 |
| Median (IQR) time from onset to ventilator, day | 10 (7.25-27) | 10 (8-17) | 10 (5-12) | .57 |

Notes: Values are numbers (percentages) unless stated otherwise. Abbreviations: N: the total number of patients with available data. IQR: interquartile range; PaO<sub>2</sub>/FiO<sub>2</sub>: ratio of partial pressure of arterial oxygen to fraction of inspired oxygen concentration; MAP: mean arterial pressure; bpm: beats per minute; CT: computerized tomography; <sup>a</sup> *P* values indicates differences between survivors and non-survivors. *P* < .05 was considered statistically significant.

Symptoms were not specific but characterized by fever (107 patients; 88.4%), dry cough (36; 52.2%), dyspnea (38; 31.4%), fatigue (33; 27.3%), sore throat and myalgia (six patients; 5.0%, respectively). Of note, ten (8.3%) cases initially presented with diarrhea or anorexia, and five (4.1%) with headache or confusion. On admission, 94 (80.3%) of 117 patients had a PaO_2_/FiO_2_ ≤ 300 mm Hg on admission, while respiratory rate maintained at 20 breaths per minute (IQR: 20-22), and heart rate at 86 beats per minute (IQR: 80-94). Shortness of breath often followed physical activities, and the time to dyspnea from the onset was 5 (IQR: 3~9) days.

#### Testing and Imaging Findings

Laboratory examination was performed and repeated on admission, day 4 (IQR: 3-6) and day 8 (IQR: 6-11) after admission, and before discharge or death (day 11; IQR: 8-16). On admission, leukopenia occurred in 33 (of 115 patients; 28.7%) patients, lymphopenia in 83 (83/115; 72.2%), thrombocytopenia in 30 (30/115; 26.1%), prolonged prothrombin time in 12 (12/101; 11.9%), hyperfibrinogenemia in 67 (67/110; 60.9%), hypoalbuminemia in 37 (37/114; 32.5%) and hyperglycemia in 58 (58/119; 48.7%), respectively. The above indicators normalized gradually on day 4 (IQR: 3-6) after admission in the survivors, while those in the non-survivors remained unchanged or worsened over time. **(Table 2)**.

**Table 2.** Laboratory findings and changes over time during hospitalization in survivors and non-survivors with SARS-CoV-2 related pneumonia.

| Laboratory findings (normal range) | On admission | Day 4 (IQR, 3-6) | Day 8 (IQR, 6-11) | On discharge or death (day 11, 8-16) | P Value<sup>a</sup> |
| --- | --- | --- | --- | --- | --- |
| Mean (SD) white blood cell, $\times 10^9/L$ (3.5-9.5) | | | | | |
| Survival | 5.5 ± 3.3 | 7.7 ± 3.6 | 8.2 ± 4.2 | 7.8 ± 4.4 | <.001 |
| < 3.5$\times 10^9/L$, n/N (%) | 28/93 (30.1) | 6/67 (9.0) | 2/50 (4.0) | 2/70 (2.9) | <.001 |
| 3.5-9.5$\times 10^9/L$, n/N (%) | 53/93 (57.0) | 44/67 (65.7) | 36/50 (72.0) | 50/70 (71.4) | .17 |
| > 9.5$\times 10^9/L$, n/N (%) | 12/93 (12.9) | 17/67 (25.4) | 12/50 (24) | 18/70 (25.7) | .13 |
| Non-survival | 6.0 ± 3.6 | 7.7 ± 3.0 | 8.5 ± 3.2 | 7.1 ± 4.1 | .19 |
| < 3.5$\times 10^9/L$, n/N (%) | 5/22 (22.7) | 0/16 (0.0) | 0/12 (0.0) | 1/18 (5.6) | .04 |
| 3.5-9.5$\times 10^9/L$, n/N (%) | 14/22 (63.6) | 14/16 (87.5) | 9/12 (75.0) | 14/18 (7.8) | .40 |
| > 9.5$\times 10^9/L$, n/N (%) | 3/22 (13.6) | 2/16 (12.5) | 3/12 (25.0) | 3/18 (16.7) | .81 |
| Mean (SD) neutrophil, $\times 10^9/L$, (1.8-6.3) | | | | | |
| Survival | 4.2 ± 3.3 | 6.3 ± 3.7 | 6.6 ± 4.1 | 5.8 ± 4.6 | <.001 |
| < 1.8$\times 10^9/L$, n/N (%) | 21/93 (22.6) | 5/67 (7.5) | 2/50 (4.0) | 4/70 (5.7) | <.001 |
| 1.8-6.3$\times 10^9/L$, n/N (%) | 53/93 (57.0) | 32/67 (47.8) | 27/50 (54.0) | 48/70 (68.6) | .10 |
| > 6.3$\times 10^9/L$, n/N (%) | 19/93 (20.4) | 30/67 (44.8) | 21/50 (42.0) | 18/70 (25.7) | .002 |
| Non-survival | 4.9 ± 3.2 | 6.5 ± 3.2 | 7.3 ± 3.4 | 5.6 ± 4.3 | .24 |
| < 1.8$\times 10^9/L$, n/N (%) | 4/22 (18.2) | 0/16 (0.0) | 0/12 (0.0) | 0/18 (0.0) | .03 |
| 1.8-6.3$\times 10^9/L$, n/N (%) | 13/22 (59.1) | 9/16 (56.3) | 4/12 (33.3) | 13/18 (72.2) | .21 |
| > 6.3$\times 10^9/L$, n/N (%) | 5/22 (22.7) | 7/16 (43.8) | 8/12 (66.7) | 5/18 (27.8) | .06 |
| Mean (SD) lymphocyte, $\times 10^9/L$, (1.1-3.2) | | | | | |
| Survival | 0.9 ± 0.4 | 1.0 ± 0.5 | 1.2 ± 0.7 | 1.4 ± 0.7 | <.001 |
| < 1.1$\times 10^9/L$, n/N (%) | 64/93 (68.8) | 44/67 (65.7) | 23/50 (46.0) | 21/70 (17.1) | <.001 |
| Non-survival | 0.7 ± 0.3 | 0.8 ± 0.5 | 0.9 ± 0.7 | 1.0 ± 0.5 | .42 |
| < 1.1$\times 10^9/L$, n/N (%) | 19/22 (86.4) | 11/16 (68.8) | 9/12 (75.0) | 11/18 (61.1) | .32 |
| Mean (SD) platelet, $\times 10^{12}/L$, (125-350) | | | | | |
| Survival | 181.5 ± 90.5 | 222.3 ± 95.3 | 232.3 ± 110.6 | 233.1 ± 103.4 | .02 |
| < 50$\times 10^{12}/L$, n/N (%) | 2/93 (2.2) | 2/67 (3.0) | 2/50 (4.0) | 2/70 (2.9) | .94 |
| 50-125$\times 10^{12}/L$, n/N (%) | 24/93 (25.8) | 8/67 (11.9) | 4/50 (8.0) | 9/70 (12.9) | .02 |
| Non-survival | 172.3 ± 66.8 | 213.7 ± 96.8 | 230 ± 123.5 | 203.9 ± 123 | .39 |
| < 50$\times 10^{12}/L$, n/N (%) | 0/22 (0.0) | 0/16 (0.0) | 0/12 (0.0) | 1/18 (5.6) | .42 |
| 50-125$\times 10^{12}/L$, n/N (%) | 4/22 (18.2) | 2/16 (12.5) | 3/12 (25.0) | 4/18 (22.2) | .84 |
| Median (IQR) AST, U/L, (0-40) |  |  |  |  |  |
| Survival | 29 (24-39) | 24 (15.3-37.8) | 25.5 (16.8-51.3) | 19.5 (15-33) | <.001 |
| > 40 U/L, n/N (%) | 20/87 (23.0) | 14/59 (23.7) | 14/42 (33.3) | 13/70 (18.6) | .36 |
| Non-survival | 40.5 (28.5-55.3) | 28.5 (17.5-47.3) | 35.5 (15.5-44.3) | 25.2(17.5-31) | .08 |
| > 40 U/L, n/N (%) | 13/26 (50.0) | 4/12 (33.3) | 4/12 (33.3) | 3/17 (17.6) | .19 |
| Median (IQR) ALT, U/L, (0-40) |  |  |  |  |  |
| Survival | 23 (15-41) | 26 (17-50) | 32.5 (23.5-74.3) | 26.5 (18.8-57.8) | .02 |
| > 40 U/L, n/N (%) | 22/87 (21.3) | 18/59 (30.5) | 18/42 (42.9) | 23/70 (32.9) | .25 |
| Non-survival | 28.5 (20.8-34.5) | 29 (21-41.5) | 31.5 (21.3-55) | 32 (21-50.5) | .74 |
| > 40 U/L, n/N (%) | 4/26 (15.4) | 3/12 (25.0) | 3/12 (25.0) | 6/17 (35.3) | .01 |
| Mean (SD) Total bilirubin, mmol/L, (3.4-20.5) |  |  |  |  |  |
| Survival | 10.3 ± 7.7 | 9.6 ± 6.7 | 10 ± 8.2 | 11.2 ± 8.4 | .66 |
| > 20.5 mmol/L, n/N (%) | 7/89 (7.9) | 5/59 (8.5) | 1/42 (2.4) | 6/70 (8.6) | .61 |
| Non-survival | 10.6 ± 6.1 | 12.4 ± 8.6 | 11.1 ± 6.7 | 13.2 ± 8.3 | .67 |
| > 20.5 mmol/L, n/N (%) | 3/26 (11.5) | 2/12 (16.7) | 1/12 (8.3) | 2/17 (11.8) | .94 |
| Mean (SD) Albumin, g/L, (34-54) |  |  |  |  |  |
| Survival | 34.9 ± 4.7 | 30.5 ± 5.7 | NA | 33.7 ± 4.9 | .09 |
| < 34 g/L, n/N (%) | 29/88 (33.0) | 12/15 (80.0) | NA | 26/75 (34.7) | .002 |
| Non-survival | 35.1 ± 4.1 | NA | NA | 31.4 ± 4.5 | .06 |
| < 34 g/L, n/N (%) | 8/26 (30.8) | NA | NA | 12/19 (63.2) | .04 |
| Mean (SD) blood glucose, mmol/L, (3.6-6.1) |  |  |  |  |  |
| Survival | 7.4 ± 4.0 | 8.1 ± 4.7 | 7.7 ± 3.6 | 6.6 ± 3.2 | .17 |
| 6.2-11.1 mmol/L, n/N (%) | 32/93 (34.4) | 30/65 (46.2) | 16/41 (39.0) | 14/66 (21.2) | .03 |
| > 11.1 mmol/L, n/N (%) | 11/93 (11.8) | 10/65 (15.4) | 7/41 (17.1) | 9/66 (13.6) | .85 |
| Non-survival | 7.5 ± 2.6 | 8.7 ± 3.5 | 10.9 ± 6.0 | 9.3 ± 6.3 | .19 |
| 6.2-11.1 mmol/L, n/N (%) | 14/26 (53.8) | 8/14 (57.1) | 4/12 (33.3) | 5/17 (29.4) | .27 |
| > 11.1 mmol/L, n/N (%) | 1/26 (3.8) | 3/14 (21.4) | 5/12 (41.7) | 5/17 (29.4) | .03 |
| Median (IQR) BUN, mmol/L, (3.1-8.0) |  |  |  |  |  |
| Survival | 4.4 (3.5-5.4) | 5.1 (3.7-7.2) | 4.7 (4.1-7.4) | 5.0 (4.1-6.3) | .01 |
| > 8.0 mmol/L, n/N (%) | 7/94 (7.4) | 9/65 (13.8) | 6/41 (14.6) | 11/67 (16.4) | .33 |
| Non-survival | 4.9 (3.6-6.4) | 5.6 (3.6-8.6) | 6.2 (3.8-7.9) | 4.9 (3.3-10.7) | .73 |
| > 8.0 mmol/L, n/N (%) | 3/26 (11.5) | 4/14 (28.6) | 2/12 (16.7) | 4/17 (23.5) | .60 |
| Median (IQR) creatinine, µmol/L, (50-120) |  |  |  |  |  |
| Survival | 71 (61-89.3) | 66 (58.5-77.5) | 62 (52.5-82.5) | 64 (53-78) | .09 |
| > 120 µmol/L, n/N (%) | 4/94 (4.3) | 2/65 (3.1) | 2/14 (14.3) | 5/67 (7.5) | .27 |
| Non-survival | 76.5 (53.8-96.3) | 67 (48.3-99) | 66.5 (46.3-99) | 65 (49-108.5) | .93 |
| > 120 µmol/L, n/N (%) | 3/26 (11.5) | 2/14 (14.3) | 1/12 (8.3) | 3/17 (17.6) | .89 |
| Mean (SD) APTT, second, (27-45) |  |  |  |  |  |
| Survival | 37.1 ± 6.1 | 32.2 ± 5.2 | 31.2 ± 6.5 | 32.3 ± 6.7 | <.001 |
| > 45 second, n/N (%) | 10/89 (11.2) | 0/32 (0.0) | 1/23 (4.3) | 1/28 (3.6) | .13 |
| Non-survival | 37.6 ± 6.1 | 32.5 ± 5.4 | 31.1 ± 3.1 | 37 ± 12.8 | .09 |
| > 45 second, n/N (%) | 3/21 (14.3) | 0/8 (0.0) | 0/7 (0.0) | 1/5 (20.0) | .82 |
| Mean (SD) prothrombin time, second, (11-16) |  |  |  |  |  |
| Survival | 14.4 ± 2.0 | 14.3 ± 2.0 | 14.5 ± 3.8 | 16.1 ± 11.7 | .43 |
| > 16 second, n/N (%) | 11/88 (12.5) | 3/32 (9.4) | 3/23 (9.4) | 3/28 (10.7) | .98 |
| Non-survival | 13.5 ± 1.7 | 16.3 ± 5.3 | 17.5 ± 9.1 | 15.1 ± 2.7 | .21 |
| > 16 second, n/N (%) | 1/21 (4.8) | 2/8 (25.0) | 1/7 (14.3) | 3/5 (60.0) | .03 |
| Mean (SD) fibrinogen, g/L, (2.0 - 4.0) |  |  |  |  |  |
| Survival | 4.2 ± 1.0 | 4.1 ± 2.2 | 3.4 ± 1.0 | 3.4 ± 1.2 | .01 |
| > 4.0 g/L, n/N (%) | 55/89 (61.8) | 16/32 (50) | 6/23 (26.1) | 8/28 (28.6) | .002 |
| Non-survival | 4.3 ± 1.1 | 3.3 ± 1.6 | 4.0 ± 2.1 | 3.7 ± 0.9 | .41 |
| > 4.0 g/L, n/N (%) | 12/21 (57.1) | 3/8 (37.5) | 3/7 (42.9) | 2/5 (40.0) | .77 |
| Positive Cardiac troponin I, n/N (%),<br>(> 0.04 ng/ml) |  |  |  |  |  |
| Survival | 10/66 (15.2) | 1/26 (3.8) | 2/15 (13.3) | 4/23 (17.4) | .48 |
| Non-survival | 1/14 (7.1) | 0/5 (0.0) | 1/3 (33.3) | 0/3 (0.0) | .49 |
| Median (IQR) procalcitonin, ng/ml, (0-0.05) |  |  |  |  |  |
| Survival | 0.07 (0.05-0.17) | 0.06 (0.04-0.09) | 0.06 (0.03-0.12) | 0.05 (0.04-0.12) | .01 |
| 0.05-0.09 ng/ml, n/N (%) | 62/87 (71.3) | 30/51 (58.8) | 18/35 (51.4) | 21/43 (48.8) | .05 |
| 1.0-1.9 ng/ml, n/N (%) | 2/87 (2.3) | 0/51 (0.0) | 0/35 (0.0) | 0/43 (0.0) | .42 |
| > 2.0 ng/ml, n/N (%) | 4/87 (4.6) | 1/51 (1.2) | 0/35 (0.0) | 2/43 (4.7) | .60 |
| Non-survival | 0.1 (0.07-0.19) | 0.08 (0.04-0.15) | 0.18 (0.09-0.5) | 0.6 (0.09-23.6) | .10 |
| 0.05-0.09 ng/ml, n/N (%) | 19/23 (82.6) | 6/9 (66.7) | 5/5 (100) | 4/6 (66.7) | .42 |
| 1.0-1.9 ng/ml, n/N (%) | 0/23 (0.0) | 0/9 (0.0) | 0/5 (0.0) | 1/6 (16.7) | .26 |
| > 2.0 ng/ml, n/N (%) | 1/23 (4.3) | 0/9 (0.0) | 0/5 (0.0) | 1/6 (16.7) | .49 |
| Mean (SD) HS-CRP, mg/L, (0-6) |  |  |  |  |  |
| Survival | 27.5 ± 10.9 | 18.5 ± 12.8 | 12.1 ± 12.6 | 10.6 ± 12.4 | <.001 |
| 6-19 mg/L, n/N (%) | 6/59 (10.2) | 10/39 (25.6) | 6/31 (19.4) | 7/40 (17.5) | .25 |
| 20-29 mg/L, n/N (%) | 13/59 (22.0) | 6/39 (15.4) | 2/31 (6.5) | 4/40 (10.0) | .17 |
| > 30 mg/L, n/N (%) | 35/59 (19.3) | 11/39 (28.2) | 6/31 (19.4) | 5/40 (12.5) | <.001 |
| Non-survival | 28.8 ± 12.8 | 23.4 ± 14.4 | 13.5 ± 14.4 | 21.1 ± 12.0 | .13 |
| 6-19 mg/L, n/N (%) | 2/18 (11.1) | 0/11 (0.0) | 0/5 (0.0) | 4/11 (36.4) | .56 |
| 20-29 mg/L, n/N (%) | 1/18 (5.6) | 2/11 (18.2) | 2/5 (40.0) | 3/11 (27.3) | .28 |
| > 30 mg/L, n/N (%) | 13/18 (72.2) | 5/11 (45.5) | 1/5 (20.0) | 3/11 (27.3) | .05 |

Notes: Values are mean (SD) unless stated otherwise. Abbreviations: N: the total number of patients with available data. IQR: interquartile range; SD: standard deviation; HS-CRP: high sensitivity of C reaction protein. AST: aspartate aminotransferase; ALT: alanine aminotransferase; APTT: activated partial thromboplastin time; BUN: blood urea nitrogen. <sup>a</sup> *P* values indicates differences over time; *P* < .05 was considered statistically significant.

The serum IgM specific for influenza A was weakly positive in one (0.9%) of 116 patients, and influenza B in three (2.6%), chlamydia and mycoplasma pneumoniae in four (3.5%), and mumps virus in one (0.9%). Candida albicans were identified on the sputum culture in two (3.3%) of 60 specimens. Procalcitonin was mildly elevated in 81 (73.6%) of 110 patients, and high sensitivity of C reaction protein elevated significantly in 70 (90.9%) of 79 patients, both of which returned gradually to normal on day 4 (IQR: 3-6) after admission in the survivors, while those in the non-survivors remained unchanged or deteriorated over time. The RT-PCR assay for SARS-CoV-2 was performed on the nasopharyngeal swab at least 2 times in 108 patients, 78 (72.2%) were positive, seven (6.5%) suspected and 23 (21.3%) negative. The time window from illness onset to a negative RT-PCR was 19 (IQR: 14-34) days in 43 cases who were initially tested positive. In 15 (13.9%) cases who had a negative RT-PCR or turned negative, retesting was positive during their hospital stay. Besides, in additional 14 patients with negative RT-PCR, 12 (85.7%) were tested positive for specific serum IgM or IgG antibody on day 28 (IQR: 10-40) after symptom onset, with a cutoff value of 2.04 (IQR: 0.18-5.13). (data available on request at the REDCap web database platform, the University of Oxford).

A total of 116 chest CT images was obtained at the early stage of the disease. The typical radiographic findings were mixed multifocal mottling (80 patients; 69%), ground-glass opacities (73; 62.9%), consolidation (18; 15.5%) and subpleural parenchymal band (8; 6.9%), distributed in the lung periphery. Bilateral lung involvement was observed in 105 patients (66.4%), and left lung in five (4.3%) and right lung in six (5.2%). Abnormalities distributed in the lung periphery were detected in 87 (75%) patients, and diffused infiltration in 22 (19%). On day 8 (IQR: 6-10) after onset, 57 chest CT scans were repeated, and 33 (57.9%) patients showed lesion absorption, 18 (31.6%) extended, six (10.5%) unchanged and 12 (21.1%) parenchymal bands. Prior to discharge or death (day 17 after onset, IQR: 12.5-22.5), 81 chest CT images were obtained, and 66 (81.5%) patients showed lesion absorption with four (5%) cleared completely, six (7.4%) enlarged, eight (9.9%) unchanged and 59 (72.8%) residual parenchymal bands. A typical chest CT finding over time was showed in **Figure 2**.

**Figure 2.**
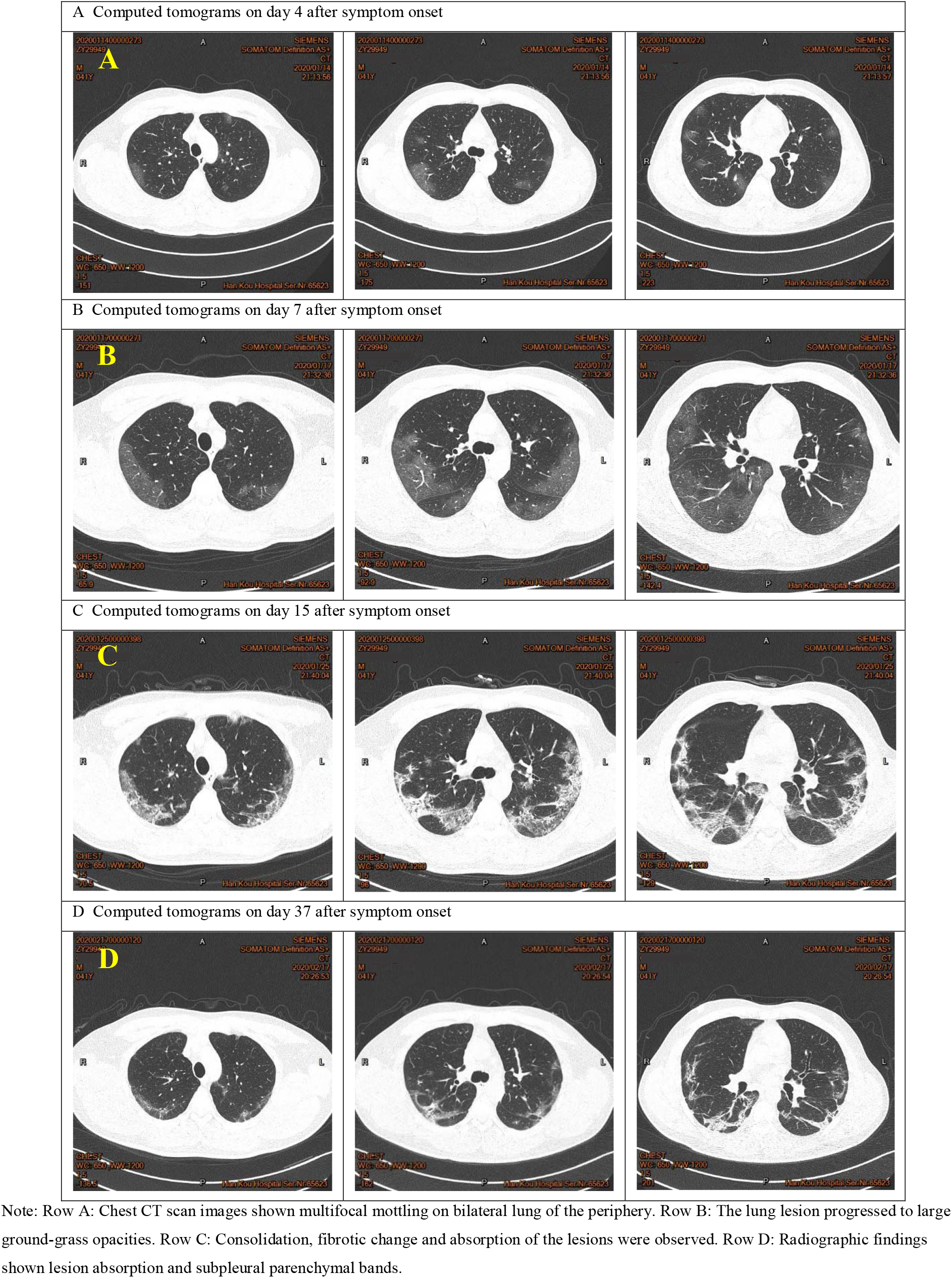
Changes in the chest computed tomograms over time in a 41 years patient with SARS-CoV-2 related pneumonia.

#### Treatment

Oseltamivir was orally administered in 79 patients (65.3%), 75 mg twice daily for five days (interquartile range 5-5.5), and Ribavirin intravenously in six (5.0%), Lopinavir/ritonavir in one (0.8%) and Chloroquine in one (0.8%) for 5-7 days, respectively. Cefoperazone/sulbactam (3 g three times a day) and Moxifloxacin (0.4 g once daily) injections were given alone or in combination in 104 (86%) for ten (interquartile range 6-12.8) days.

Piperacillin/tazobactam, Meropenem, Vancomycin and linezolid were also used alone or in combination in 34 (28.1%) patients. In addition, Methylprednisolone (0.5-1 mg/kg, once or twice daily) was given intravenously and taped in 100 cases (82.7%) for 10 days (IQR: 5.25-12), and Immunoglobulin (10 g, once daily, intra-venously) in 71 (58.7%) for six days (IQR: 3-9). Moreover, Thymopentin (10 mg, 3 times a week, subcutaneously) and Cordyceps Sinensis (herbal medicine, orally) were also prescribed in patients with lymphopenia.

Nasal cannula oxygen was required in 68 (56.2%) patients, oxygen mask with a reservoir in 16 (13.2%), and high flow nasal cannula in four (3.2%). Noninvasive ventilation was implemented in 17 (14.1%) critically ill patients for seven days (IQR: 1.75-10.5), and six of whom died. Invasive ventilation was instituted in three patients (11.5%), and tracheotomy performed in one case, all of the three patients died in 2, 11 and 12 days (**Table 3**). The time from onset to mechanical ventilation was ten (IQR: 7.3-14) days.

**Table 3.**
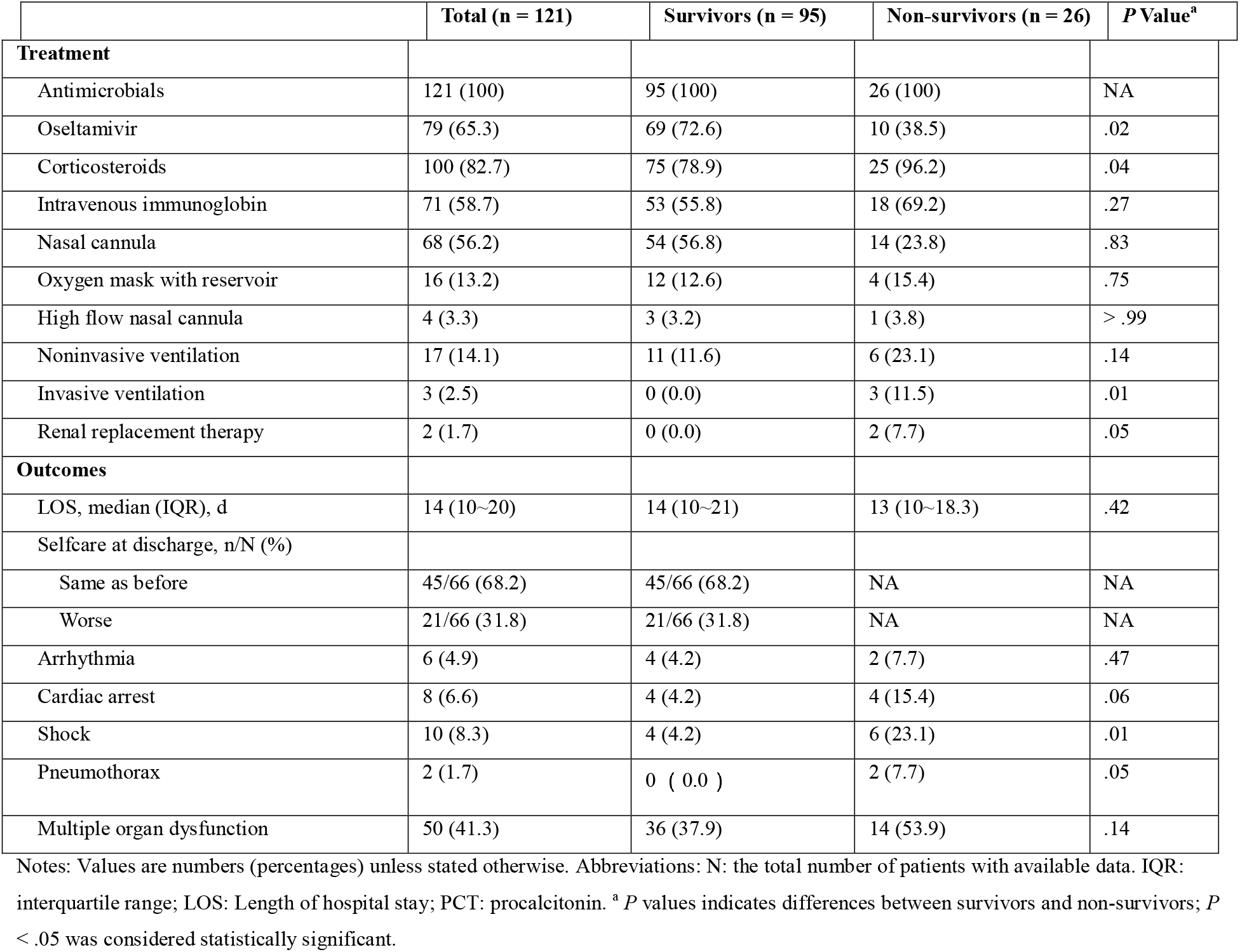
| Treatment and outcomes in survivors and non-survivors with SARS-CoV-2 related pneumonia.

|  | Total (n = 121) | Survivors (n = 95) | Non-survivors (n = 26) | P Value<sup>a</sup> |
| --- | --- | --- | --- | --- |
| **Treatment** |  |  |  |  |
| Antimicrobials | 121 (100) | 95 (100) | 26 (100) | NA |
| Oseltamivir | 79 (65.3) | 69 (72.6) | 10 (38.5) | .02 |
| Corticosteroids | 100 (82.7) | 75 (78.9) | 25 (96.2) | .04 |
| Intravenous immunoglobulin | 71 (58.7) | 53 (55.8) | 18 (69.2) | .27 |
| Nasal cannula | 68 (56.2) | 54 (56.8) | 14 (23.8) | .83 |
| Oxygen mask with reservoir | 16 (13.2) | 12 (12.6) | 4 (15.4) | .75 |
| High flow nasal cannula | 4 (3.3) | 3 (3.2) | 1 (3.8) | > .99 |
| Noninvasive ventilation | 17 (14.1) | 11 (11.6) | 6 (23.1) | .14 |
| Invasive ventilation | 3 (2.5) | 0 (0.0) | 3 (11.5) | .01 |
| Renal replacement therapy | 2 (1.7) | 0 (0.0) | 2 (7.7) | .05 |
| **Outcomes** |  |  |  |  |
| LOS, median (IQR), d | 14 (10~20) | 14 (10~21) | 13 (10~18.3) | .42 |
| Selfcare at discharge, n/N (%) |  |  |  |  |
| Same as before | 45/66 (68.2) | 45/66 (68.2) | NA | NA |
| Worse | 21/66 (31.8) | 21/66 (31.8) | NA | NA |
| Arrhythmia | 6 (4.9) | 4 (4.2) | 2 (7.7) | .47 |
| Cardiac arrest | 8 (6.6) | 4 (4.2) | 4 (15.4) | .06 |
| Shock | 10 (8.3) | 4 (4.2) | 6 (23.1) | .01 |
| Pneumothorax | 2 (1.7) | 0 ( 0.0 ) | 2 (7.7) | .05 |
| Multiple organ dysfunction | 50 (41.3) | 36 (37.9) | 14 (53.9) | .14 |

Notes: Values are numbers (percentages) unless stated otherwise. Abbreviations: N: the total number of patients with available data. IQR:
interquartile range; LOS: Length of hospital stay; PCT: procalcitonin. <sup>a</sup> P values indicates differences between survivors and non-survivors; P < .05 was considered statistically significant.

#### Complications and outcomes

Of 121 patients, 23 (19.0%) were moderately, 41 (33.9%) severely and 57 (47.1%) critically ill. Hypoxemia occurred in 94 (94/117; 80.3%) patients, multiple organ dysfunction in 50 (41.3%), shock in ten (8.3%) and cardiac arrest in eight (6.6%). At the end point of the study, 95 (78.5%) patients discharged alive, and 26 (21.5%) dead. The case fatality rates were 4.4% (one of 23 patients), 29.3% (12/41), 22.8% (13/57) or 45% (9/20) for patients with moderate, severe, critical illness or requiring invasive and non-invasive ventilation. The length of hospital stay was 14 (IQR: 10-20) days, and selfcare ability worsened in 21 patients (21/66; 31.8%) cases. The Logistic regression analysis showed that the age and blood albumin level were the independent risk factors for death, with odds ratios of 1.18 (95% CI, 1.04-1.32, *P* < .01) *vs* 1.35 (95% CI, 1.0-1.8, *P* < .05), respectively. Kaplan-Meier curves of cumulative survival probability showed that patients over 60 years old were most likely to have poorer outcomes (**Figure 3**).

**Figure 3.**
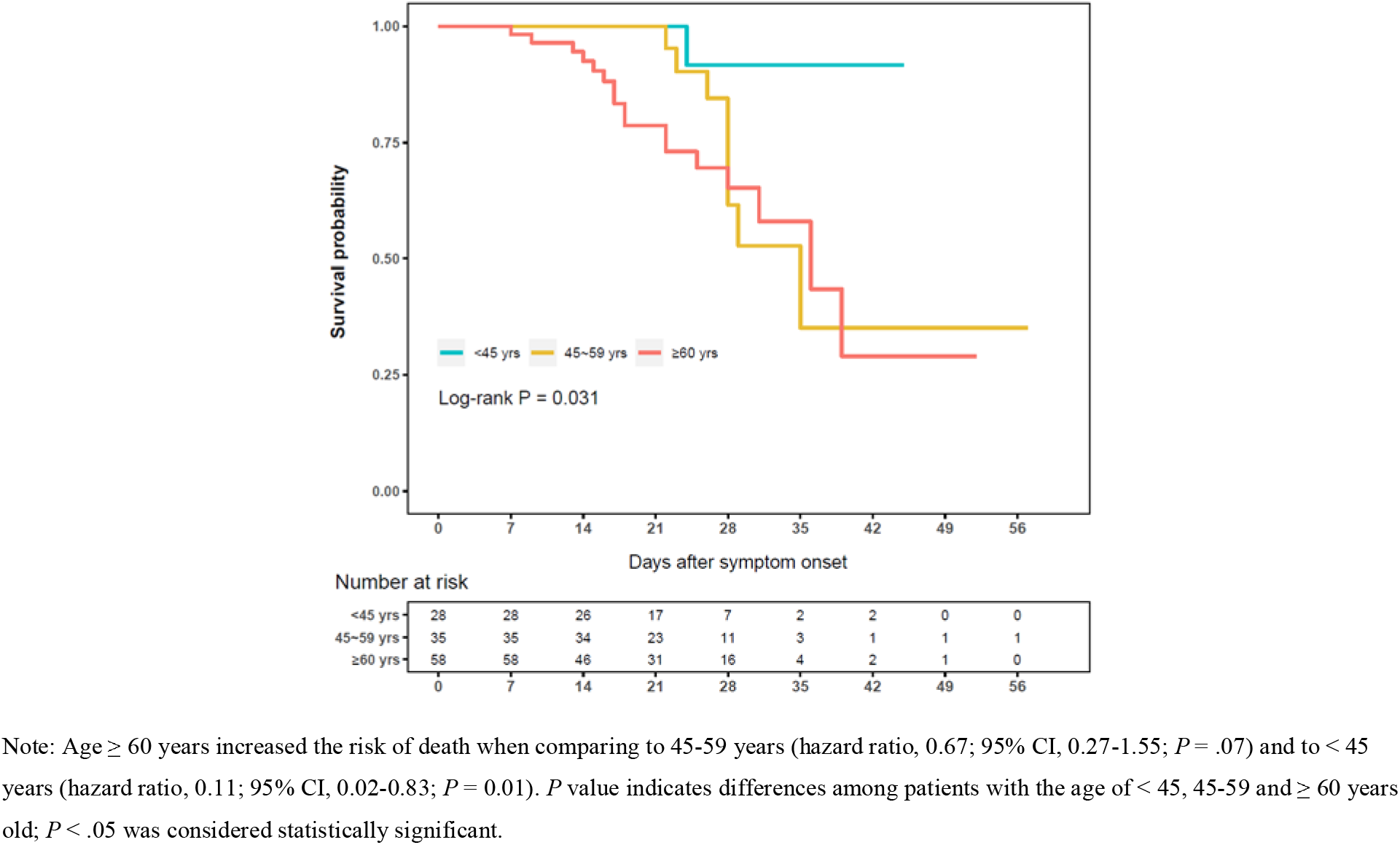
Kaplan-Meier curves of cumulative survival probability for patients with SARS-CoV-2 related pneumonia and with the age of < 45, 45-59 and ≥ 60 years on day 56 after symptom onset.

### Discussion

In this single-centered retrospective observational cohort study, we reported the case fatality rate and natural course of 121 patients with laboratory-confirmed or suspected SARS-CoV-2 pneumonia, while specific drug for COVID-19 was currently not available.^1^ In contrast to the first-generation of patients,^7^ near half of whom had a history of exposure to the Seafood wholesale market, most of our patients did not present definite epidemiological factors, except for 12 (9.9%) had exposure to healthcare facilities, 9 (7.4%) closely contacted to COVID-19 patients, 6 (5%) were familial clusters and 2 (1.7%) doctors. Given that human-to-human and hospital or work-related transmission had been presumed^10, 22, 32^, they might be representative of the first and next generation of infections in the early stage of the epidemic in Wuhan, China. Of these patients, 23 (19.0%) were moderately, 41 (33.9%) severely and 57 (47.1%) critically ill. Ninety-five patients (78.5%) discharged alive, and 26 (21.5%) died in-hospital. The case fatality rates were 4.4%, 29.3%, 22.8% or 45% for patients with moderate, severe, critical illness or requiring invasive and non-invasive ventilation. Age and blood albumin level were identified as the independent risk factors for death, and patients over 60 years old were most likely to have poorer outcomes.

#### Interpretation

The reason why we enrolled patients without selectivity, either with suspected or confirmed SARS -CoV-2 related pneumonia, was that it might well reflect the disease course, recovery and fatality in this subset of patients with varied severities of illness. To our patients, negative RT-PCR could not completely rule out SARS-CoV-2 related pneumonia. ^23 24^ Firstly, RT-PCR testing was not available for our patients until in the late stage of the epidemic, and patients admitted before that time might be misdiagnosed. In 14 additional patients who were treated contemporarily in our ward and remained negative RT-PCR, serum IgM or IgG antibody specific for SARS-CoV-2 was tested positive in 12 (85.7%) cases on day 28 (interquartile range 10-40) after admission, meaning that they might have been infected with SARS-CoV-2 within weeks. Secondly, false-negative RT-PCR for SARS-CoV-2 had been reported.^23 24^ Xu and co-workers reported that false-negative RT-PCR occurred in 691 (52.2%) of 1,324 outpatients with COVID-19 infection in their first test.^23^ Xie and investigators reported five patients who had typical clinical and radiological manifestations but initial RT-PCR negative, retesting was positive 2-7 days after the first chest CT scan.^24^ The authors argued that a chest CT image might be more sensitive to the diagnosis of SARS-CoV-2 related pneumonia. Our data showed that 15 patients (11.6%) whose initial RT-PCR were negative, retesting was positive during their hospital stay. Reasons for false-negative RT-PCR might be attributed to the sensitivity of the diagnostic test kits, inappropriate sampling and insufficient viral load of the specimens. Repeated tests or newly developed detection methods should be adopted to improve diagnostic efficiency.^24^ Thirdly, our data shown that the time window of RT-PCR assay for SARS-CoV-2 was 19 (interquartile range 14-34) days from illness onset, the viral load on nasal and throat swabs would be too trace to be detected after 18 days from onset.^25^ Finally, all of our patients presented very similar epidemiological, clinical and radiographic features, and other probable causative agents was excluded, except 9 (7.8%) of 116 cases who shown weakly positive of serum IgM antibodies for influenza A or B, chlamydia or mycoplasma pneumoniae and mumps virus. In addition, of the 60 sputum samples, candida albicans were detected in 2 (3.3%), but no bacteria were identified.

The SARS-CoV-2 infection had exhibited somewhat the property of self-limiting disease. The treatment for our patients was largely supportive, except a few received Ribavirin or Chloroquine, while absorption of the lesions on chest CT images could be observed in 57.9% of patients on day 8 after onset, along with a remarkable increase in lymphocyte count in the survivors. Pan and coworkers reported 21 cases with SARS-CoV-2 pneumonia, the greatest lung involvement was achieved on day 10 after symptom onset, and the lesions gradually absorbed after 2 weeks.^26^ Of note, an obvious discrepancy between symptom and the degree of hypoxemia was found in a considerable number of our patients, 94 of 117 patients (80.3%) had a PaO_2_/FiO_2_ of less than 300 mm Hg on admission, but they did not present obvious dyspnea or tachycardia at rest, and the extent of lung involvement on their chest CT image was not always consistent with the degree of hypoxemia. This so-called “atypical ARDS” could be explained by the autopsy findings in non-mechanically ventilated patients with confirmed SARS-CoV-2 pneumonia.^28-30^ The pulmonary pathology of SARS-CoV-2 related pneumonia greatly resembled those of SARS-CoV and Middle Eastern respiratory syndrome (MERS) coronavirus infections, but the severity degree of the pulmonary edema, hyaline membrane formation and vasculopathy were less apparent in the early or acute exudation stage. This autopsy findings were consistent with the chest CT imaging features, on which the pulmonary edema or pleural effusion was rarely observed. In our study, patients mostly died of acute exacerbation of hypoxemic respiratory failure rather than multiple organ dysfunction, the “atypical ARDS” might represent a plateau in the course of the disease, after which the disease might worsen or improve. It was crucial to maintain the pulse oxygen saturation at an acceptable level through aggressive oxygen support, e.g., ≥93%, thus to prevent patients from developing severe respiratory distress.^27^ If the patients could get through the acute phase of the illness, they were more likely to recover. Besides, Li and co-authors presumed that SARS-CoV-2 might have potential neuroinvasion by transsynaptic transfer, and be partially related to the aggravation of respiratory failure in the COVID-19 patients. Early use of inhaled antiviral agents might be useful to inhibit the SARS-CoV-2 replication in the respiratory tracts and lung, and to prevent from its subsequent neuroinvasion.^31^

Mechanical ventilation or extracorporeal life support seemed less effective on patients with refractory hypoxemic respiratory failure. In our study, invasive or noninvasive ventilation was adopted in 20 critically ill patients, nine (45%) of them died in 12 days. Zhou and colleagues analyzed retrospectively 191 cases with a median age of 56 years old, twenty-six patients had received non-invasive ventilation and thirty-two invasive ventilation, but fifty-five (94.8%) of whom died. ECMO therapy was used for three cases but all died.^33^ Huang and co-investigators reported 41 cases who were confirmed as SARS-CoV-2 pneumonia and admitted in the earliest stage of the outbreak, invasive or non-invasive ventilation was required in ten of 13 critically ill patients, five (39%) of them died.^7^ Although the case fatality rate of critically ill patient was lower than ours, their patients were younger, with a median age of 49 (interquartile range 41-58) years *vs* 59 (interquartile range 46-67) years, respectively. As of the final date of follow-up, seven cases were still hospitalized, and the case fatality rate was expected to increase. Yang and co-workers reported 52 critically ill cases, with an average age of 59.7 years old.^11^ Mechanical ventilation was instituted in 37 cases, and 30 (81%) of whom died in 28 days. ECMO therapy was given in 6 patients, five of them died and one still on the machine as of day 28. The authors explained that the case fatality rate of SARS-CoV-2 infection was likely to be higher in critically ill patients than those of SARS (26%-43%), MERS (58%) and severe ARDS (50%).^11^ Patients who were male sex, older (> 65 years) and complicated with the underlying disease were more likely to develop critical illness and death. The reason why mechanical ventilation and ECMO therapy were less effective on ARDS caused by SARS-CoV-2 might be explained by the pathological changes from a COVID-19 patient.^36^ In mechanically ventilated patient, multiple organ dysfunction was common,^13^ and the lung was more severely injured than previously described.^28-30^ In the hemorrhagic area, the pulmonary pathology showed “alveolar tamponade”, consisted of extensive fibrinoid exudates, together with a large number of red blood cell, type II alveolar epithelial cells, macrophages and tissue cells. In the non-hemorrhage area, massive “thick mucus embolus” plugged the dilated bronchioles and terminal bronchial lumens, and the fibrinoid exudates deposited on the walls of pulmonary small vessels and formed ground-glass appearance, causing stenosis and occlusion of the vascular lumen. This “vasculopathy” was different from that of “typical ARDS”, which mainly manifested with “capillary leakage”.^18^ In a correspondence,^34^ Henry proposed that ECMO therapy might induce potential harms to the COVID-19 patients by substantially reducing in the number and function of some populations of lymphocytes and elevating the serum level of IL-6 concentrations. The repletion of lymphocytes could be key to recovery from the COVID-19, and IL-6 have a potential damage to the lung parenchyma. When selecting candidates for ECMO, the immunological status of the patients should be considered.

Steroids treatment for SARS-CoV-2 pneumonia with ARDS was controversial. Some believed that glucocorticoids had no mortality benefit in patients with SARS or MERS, but delayed viral clearance and even sped up the viral replication in neurons.^31 35^ Others argued that the serum levels of cytokines were higher in critically ill patients infected with SARS-CoV-2, with over-activation of proinflammatory and cytotoxic T cells, and appropriate use of steroid could reduce the inflammatory-induced lung injury.^10 28^ In a case-control study, Wu and co-authors found that methylprednisolone decreased the risk of death in patients with SARS-CoV-2 and ARDS (HR, 0.38; 95% CI, 0.20-0.72).^14^ In our study, methylprednisolone was given intravenously and taped in 82.7% of patients for 10 days, with a dosage of 0.5-1 mg/kg. Although the time for viral clearance, lesion absorption on chest CT image and hospital stay did not differ significantly from those reported in the literature,^7 10-12 25 33^ statistical analysis showed the use of corticosteroids might be a risk factor for death. Besides, Thymopentin, Cordyceps Sinensis (herbal medicine) and Immunoglobulin were prescribed to our patients with lymphopenia, and increases in blood lymphocyte count were observed in 4 days, along with the remission of the disease.

#### Limitations

There are several limitations in this study. First, in our patient cohort, 78 patients (64.5%) were confirmed to have SARS-CoV-2 pneumonia, while 43 (35.5%) were suspected. Although no other probable etiologies were identified, it might still impose an influence on the reliability of the study. However, we tested 14 patients who had a negative RT-PCR, 12 (85.7%) were positive for serum IgM or IgG antibody specific for SARS-CoV-2, highly suggesting that they had been infected with SARS-CoV-2 within weeks. Therefore, more sensitive and novel detection methods should be adopted to improve the diagnostic efficiency. Second, sample size calculation was waived in our study, and the number of patients for each subset was small, thus the case fatality rates calculated might not reflect the actual deaths of the patients. Data from other studies on SARS-CoV-2 pneumonia were needed to provide a case-fatality spectrum in patients with varied severities of illness. Third, for this single-center, retrospective cohort study, there might be existing potential sources of bias in selection of participants, groupings, comparability between the survivors and non-survivors, and data missing due to incomplete medical records, etc., all of these factors might have an impact on the power of statistics. The statistical results should be interpreted with caution, and large-scaled, prospective cohort studies were needed to justify the findings of this study in the future.

## CONCLUSION

Although the COVID-19 had shown the property of self-limiting, the case fatality rate in-hospital was high in severely or critically ill patients with SARS-CoV-2 related pneumonia. Hence, for the elderly patient with hypertension, close monitoring and appropriate supportive treatment should be taken earlier and aggressively to prevent from developing severe or critical illness. Corticosteroid use might link to death. Repeated RT-PCR tests or novel detection methods for SARS-CoV-2 should be adopted to improve diagnostic efficiency.

## Data Availability

The data that support the findings of this study will be entered online and deposited on the International Severe Acute Respiratory and Emerging Infection Consortium Data Platform (ISARIC DP) at https://ncov.medsci.ox.ac.uk/, hosted by the University of Oxford. The data will be made available on reasonable requests to the corresponding author. A proposal with detailed description of study objectives and statistical analysis plan would be needed for evaluation of the reasonability of requests. Additional materials might also be required during the process of evaluation. Participant data in the form of pseudonymization will be provided after approval from the corresponding author and Wuhan Hankou Hospital.

## Contributors

HW, YL, QL, XW, TH, and KW conceived and designed this study. YL, XW, TH, KW, and YLiu collected, analyzed and interpreted the data. QL, YH, LY, and HF provided administrative, technical, or material support. HW, ZB, and LX performed the statistical analysis. HW drafted the manuscript. HW and QL modified the final manuscript. HW had full access to all of the data in the study and takes responsibility for the integrity of the data and the accuracy of the data analysis. YL and QL contributed equally and share first authorship. XW, TH and KW contributed equally to this article. All authors listed meet authorship criteria and no others have been omitted. HW is the guarantor.

## Funding

This work was supported by the Clinical Research Startup Program of Southern Medical University by High-level University Construction Funding of Guangdong Provincial Department of Education (grant number: LC2016PY036 to Dr Wang). The funders had no role in the design and conduct of the study; collection, management, analysis, and interpretation of the data; preparation, review, or approval of the manuscript; and decision to submit the manuscript for publication.

## Competing interest

All authors have completed the Unified Competing Interest form. No support from any organization for the submitted work; no financial relationships with any organizations that might have an interest in the submitted work in the previous three years, no other relationships or activities that could appear to have influenced the submitted work.

## Patient and public involvement

Patients and/or the public were not involved in the design, or conduct, or reporting, or dissemination plans of this research.

## Patient consent for publication

Not required.

## Ethics approval

Ethics approval was obtained from the Hankou Hospital Medical Research Ethics Committee (No. HKYY-2020-028), and written informed consent was waived for this retrospective study.

## Transparency declaration

The lead author affirms that the manuscript is an honest, accurate, and transparent account of the study being reported; that no important aspects of the study have been omitted; and that any discrepancies from the study as planned have been explained.

## Open access

This is an open access article distributed in accordance with the Creative Commons Attribution 4.0 Unported (CC BY 4.0) license, which permits others to copy, redistribute, remix, transform and build upon this work for any purpose, provided the original work is properly cited, a link to the license is given, and indication of whether changes were made. See: https://creativecommons.org/licenses/by/4.0/.

## Acknowledgement

The authors thank all patients and medical staff of Wuhan Hankou Hospital, and pay tribute to healthcare workers who have sacrificed their lives or are currently working on the frontline of the epidemic. We thank Prof. Mengfeng Li, Department of Administration, Southern Medical University, for critical revision and professional advice on this study. We also thank Chong-yang Duan, MD, Department of Biostatistics, School of Public Health, Southern Medical University for support and work on data processing and statistical analysis.

## ORCID iDs

Hua Wang http://orcid.org/0000-0002-2869-9488

## Exclusive Licence

I, the Submitting Author has the right to grant and does grant on behalf of all authors of the Work (as defined in the below author licence), an exclusive licence and/or a non-exclusive licence for contributions from authors who are: i) UK Crown employees; ii) where BMJ has agreed a CC-BY licence shall apply, and/or iii) in accordance with the terms applicable for US Federal Government officers or employees acting as part of their official duties; on a worldwide, perpetual, irrevocable, royalty-free basis to BMJ Publishing Group Ltd (“BMJ”) its licensees and where the relevant Journal is co-owned by BMJ to the co-owners of the Journal, to publish the Work in BMJ Open and any other BMJ products and to exploit all rights, as set out in our licence.

The Submitting Author accepts and understands that any supply made under these terms is made by BMJ to the Submitting Author unless you are acting as an employee on behalf of your employer or a postgraduate student of an affiliated institution which is paying any applicable article publishing charge (“APC”) for Open Access articles. Where the Submitting Author wishes to make the Work available on an Open Access basis (and intends to pay the relevant APC), the terms of reuse of such Open Access shall be governed by a Creative Commons licence – details of these licences and which Creative Commons licence will apply to this Work are set out in our licence referred to above.

## Notes

### Competing Interest Statement

The authors have declared no competing interest.

